# At what times during infection is SARS-CoV-2 detectable and no longer detectable using RT-PCR based tests?: A systematic review of individual participant data

**DOI:** 10.1101/2020.07.13.20152793

**Authors:** Sue Mallett, A. Joy Allen, Sara Graziadio, Stuart Taylor, Naomi S Sakai, Kile Green, Jana Suklan, Chris Hyde, Bethany Shinkins, Zhivko Zhelev, Jaime Peters, Philip Turner, Nia W. Roberts, Lavinia Ferrante di Ruffano, Robert Wolff, Penny Whiting, Amanda Winter, Gauraang Bhatnagar, Brian D. Nicholson, Steve Halligan

## Abstract

**Background:** Tests for severe acute respiratory syndrome coronavirus 2 (SARS-CoV-2) viral ribonucleic acid (RNA), using reverse transcription polymerase chain reaction (RT-PCR) are pivotal to detecting current coronavirus disease (COVID-19) and duration of detectable virus indicating potential for infectivity.

**Methods:** We conducted an individual participant data (IPD) systematic review of longitudinal studies of RT-PCR test results in symptomatic SARS-CoV-2. We searched PubMed, LitCOVID, medRxiv and COVID-19 Living Evidence databases. We assessed risk of bias using a QUADAS- 2 adaptation. Outcomes were the percentage of positive test results by time and the duration of detectable virus, by anatomical sampling sites.

**Findings:** Of 5078 studies screened, we included 32 studies with 1023 SARS-CoV-2 infected participants and 1619 test results, from -6 to 66 days post-symptom onset and hospitalisation. The highest percentage virus detection was from nasopharyngeal sampling between 0 to 4 days post-symptom onset at 89% (95% confidence interval (CI) 83 to 93) dropping to 54% (95% CI 47 to 61) after 10 to 14 days. On average, duration of detectable virus was longer with lower respiratory tract (LRT) sampling than upper respiratory tract (URT). Duration of faecal and respiratory tract virus detection varied greatly within individual participants. In some participants, virus was still detectable at 46 days post-symptom onset.

**Interpretation:** RT-PCR misses detection of people with SARS-CoV-2 infection; early sampling minimises false negative diagnoses. Beyond ten days post-symptom onset, lower RT or faecal testing may be preferred sampling sites. The included studies are open to substantial risk of bias so the positivity rates are probably overestimated.

**PANEL: RESEARCH IN CONTEXT:** *Evidence before this study:* There are numerous reports of negative severe acute respiratory syndrome coronavirus 2 (SARS-CoV-2) reverse transcription polymerase chain reaction (RT-PCR) test results in participants with known SARS-CoV-2 infection, and increasing awareness that the ability of RT-PCR tests to detect virus depends on the timing of sample retrieval and anatomical sampling site. Individual studies suggest that positive test results from RT-PCR with nasopharyngeal sampling declines within a week of symptoms and that a positive test later in the disease course is more likely from sputum, bronchoalveolar lavage (BAL) or stool, but data are inconsistent.

*Added value of this study:* We searched 5078 titles and abstracts for longitudinal studies reporting individual participant data (IPD) for RT-PCR for participants with COVID-19 linked to either time since symptom onset or time since hospitalisation. Search included SARS-CoV-2 and RT-PCR keywords and MeSH terms. Each included study was subject to careful assessment of risk of bias. This IPD systematic review (SR) addresses RT-PCR test detection rates at different times since symptom onset and hospitalisation for different sampling sites, and summarises the duration of detectable virus. To our knowledge, this is the first rapid SR addressing this topic. We identified 32 studies available as published articles or pre-prints between January 1^st^ and April 24^th^ 2020, including participants sampled at 11 different sampling sites and some participants sampled at more than one site. At earlier time points, nasopharyngeal sampling had the highest virus detection, but the duration of shedding was shorter compared to lower respiratory tract sampling. At 10 to 14 days post-symptom onset, the percentage of positive nasopharyngeal test results was 54% compared to 89% at day 0 to 4. Presence and duration of faecal detection varied by participant, and in nearly half duration was shorter than respiratory sample detection. Virus detection varies for participants and can continue to be detected up to 46 days post-symptom onset or hospitalisation. The included studies were open to substantial risk of bias, so the detection rates are probably overestimates. There was also poor reporting of sampling methods and sparse data on sampling methods that are becoming more widely implemented, such as self-sampling and short nasal swab sampling (anterior nares/mid turbinate).

*Implications of all the available evidence:* Results from this IPD SR of SARS-CoV-2 testing at different time points and using different anatomical sample sites are important to inform strategies of testing. For prevention of ongoing transmission of SARS-CoV-2, samples for RT-PCR testing need to be taken as soon as possible post-symptom onset, as we confirm that RT-PCR misses more people with infection if sampling is delayed. The percentage of positive RT-PCR tests is also highly dependent on the anatomical site sampled in infected people. Sampling at more than one anatomical site may be advisable as there is variation between individuals in the sites that are infected, as well as the timing of SARS-CoV-2 virus detection at an anatomical site. Testing ten days after symptom onset will lead to a higher frequency of negative tests, particularly if using only upper respiratory tract sampling. However, our estimates may considerably understate the frequency of negative RT-PCR results in people with SARS-CoV- 2 infection. Further investment in this IPD approach is recommended as the amount data available was small given the scale of the pandemic and the importance of the question. More studies, learning from our observations about risk of bias and strengths of example studies (Box 1, Box 2) are urgently needed to inform the optimal sampling strategy by including self-collected samples such as saliva and short nasal swabs. Better reporting of anatomical sampling sites with a detailed methodology on sample collection is also urgently needed.

## MAIN TEXT

### INTRODUCTION

Accurate testing is pivotal to controlling severe acute respiratory syndrome coronavirus 2 (SARS-CoV-2), otherwise known as the coronavirus disease 2019 (COVID-19). Considerable political and medical emphasis has been placed on rapid access to testing both to identify infected individuals so as to direct appropriate therapy, appropriate return to work, and to implement containment measures to limit the spread of disease. However, success depends heavily on test accuracy. Understanding when in the disease course the virus is detectable is important for two purposes, firstly to understand when and how to detect SARS-CoV-2, and secondly to understand how long individuals are likely to remain infective posing a risk to others.

The success of COVID-19 testing depends heavily on the use of accurate tests at the appropriate time. Testing for active virus infection relies predominantly on reverse transcription polymerase chain reaction (RT-PCR), which detects viral ribonucleic acid (RNA) that is shed in varying amounts from different anatomical sites and at different times during the disease course. It is increasingly understood that differences in virus load impact directly on diagnostic accuracy, notably giving rise to negative tests in disease-positive individuals^1^. Positivity is contingent upon sufficient virus being present to trigger a positive test which may depend on test site, sampling methods and timing^2^. For example, it is believed that positive nasopharyngeal RT-PCR declines within a week of symptoms so that a positive test later in the disease course is more likely from sputum, bronchoalveolar lavage fluid or stool^3^. Nomenclature for anatomical site is also unclear, with a wide variety of overlapping terms used such as “oral”, “throat”, “nasal”, “pharyngeal”, “nasopharyngeal”.

Because testing is pivotal to management and containment of COVID-19, we performed an individual participant data (IPD) systematic review of emerging evidence about test accuracy by anatomical sampling site to inform optimal sampling strategies for SARS-CoV-2. We aimed to examine at what time points during SARS-CoV-2 infection it is detectable at different anatomical sites using RT-PCR based tests.

## METHODS

This IPD systematic review followed the recommendations of the PRISMA-IPD checklist^4^.

### Eligibility

Eligible articles were any case-series or longitudinal studies reporting participants with confirmed COVID-19 tested at multiple times during their infection and provided IPD for RT-PCR test results at these times. We stipulated that test timings were linked to index dates of time since symptom onset or time since hospital admission as well as COVID-19 diagnosis by positive RT-PCR and/or suggestive clinical criteria, for example World Health Organization (WHO) guidelines^5^.

### Search strategy and article selection

Search strings were designed and conducted subsequently in PubMed, LitCOVID and medRxiv by an experienced information specialist (NR). The search end date was 24^th^ April 2020. We additionally included references identified by COVID-19: National Institute for Health Research (NIHR) living map of living evidence (http://eppi.ioe.ac.uk/COVID19_MAP/covid_map_v4.html), COVID-19 Living Evidence (https://ispmbern.github.io/covid-19/living-review/) with a volunteer citizen science team, “The Virus Bashers”. Additional details S1.

### Data extraction

Data were extracted into pre-specified forms. We did not contact authors for additional information. Study, participant characteristics, and ROB were extracted in Microsoft Excel (KG, JS, SG, JA, AW, SM). Data included country, setting, date, number of participants and IPD participants, inclusion criteria, IPD selection, participant age, sample types, RT-PCR test type and equipment, primers. RT-PCR test results were extracted using Microsoft Access (SM, BS, JP, ZZ, CH).

### Risk of Bias

We could not identify an ideal risk-of-bias (ROB) tool for longitudinal studies of diagnostic tests, so we adapted the risk of bias tool for diagnostic accuracy studies QUADAS-2^6^ to include additional signalling questions to cover anticipated issues. ROB signalling questions, evaluation criteria and domain assessment of potential bias are reported (S2).

### Sampling method and grouping

Details of sampling sites and methods, including location of the sampling site(s), any sample grouping (for example, if combined throat and nasal swabs), were extracted from full texts by a clinician (NS) with queries referred to a second clinician (ST). If stated, details of sampling methodology were recorded, including who collected samples, information regarding anatomical location (e.g. how the nasopharynx was identified), and sample storage. Additional details S3.

### RT-PCR test result conversion to binary results

IPD RT-PCR results were extracted from each article and converted to binary results (“positive” or “negative”). Data from Kaplan-Meier (KM) curves were extracted using Web digitizer ^7^. Additional details S3.

### Data analysis

Days since symptom onset and days since hospital admission were calculated from reported IPD. Data were presented collated across 5-day time intervals for each sample method, with longer times grouped within the longest time interval, and 95% CI were calculated for proportions. For comparison of duration of positive RT-PCR from respiratory tract (RT) and faecal samples, analysis and graphical presentation was restricted to participants sampled by both methods. Data analysis used STATA (14.2 StataCorp LP, Texas, USA). Additional details S3.

## RESULTS

### Included studies

5078 articles were identified, 116 full text articles were screened, and 32 articles were included^8-39^ (Figure 1). Most articles were from China, in hospitalised adults participants (Table 1). Articles reported on a total of 1023 participants and 1619 test results. Twenty-six (81%) articles reported data on test results since the start of symptoms, 23 (72%) since hospital admission. Sixteen studies including 22% (229/1023) of the participants reported both these time points: The median time between symptom onset and hospitalisation was 5 days (interquartile range (IQR) 2 to 7 days). The median number of participants per study was 22 (IQR 9 to 56, range 5 to 232) and the median number of RT-PCR test results per participant was 4 (IQR 2 to 9) (Table 2).

**Table 1:**
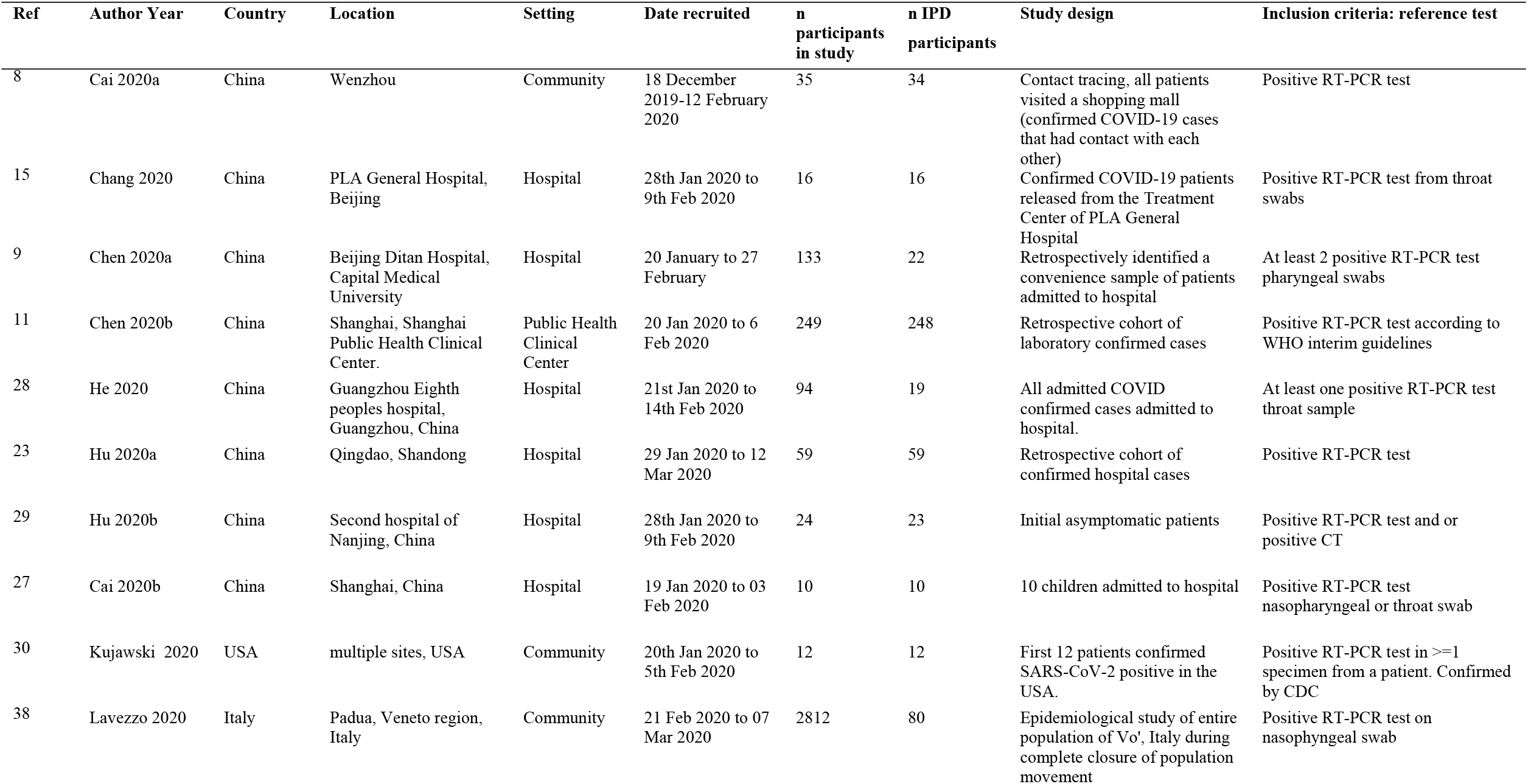
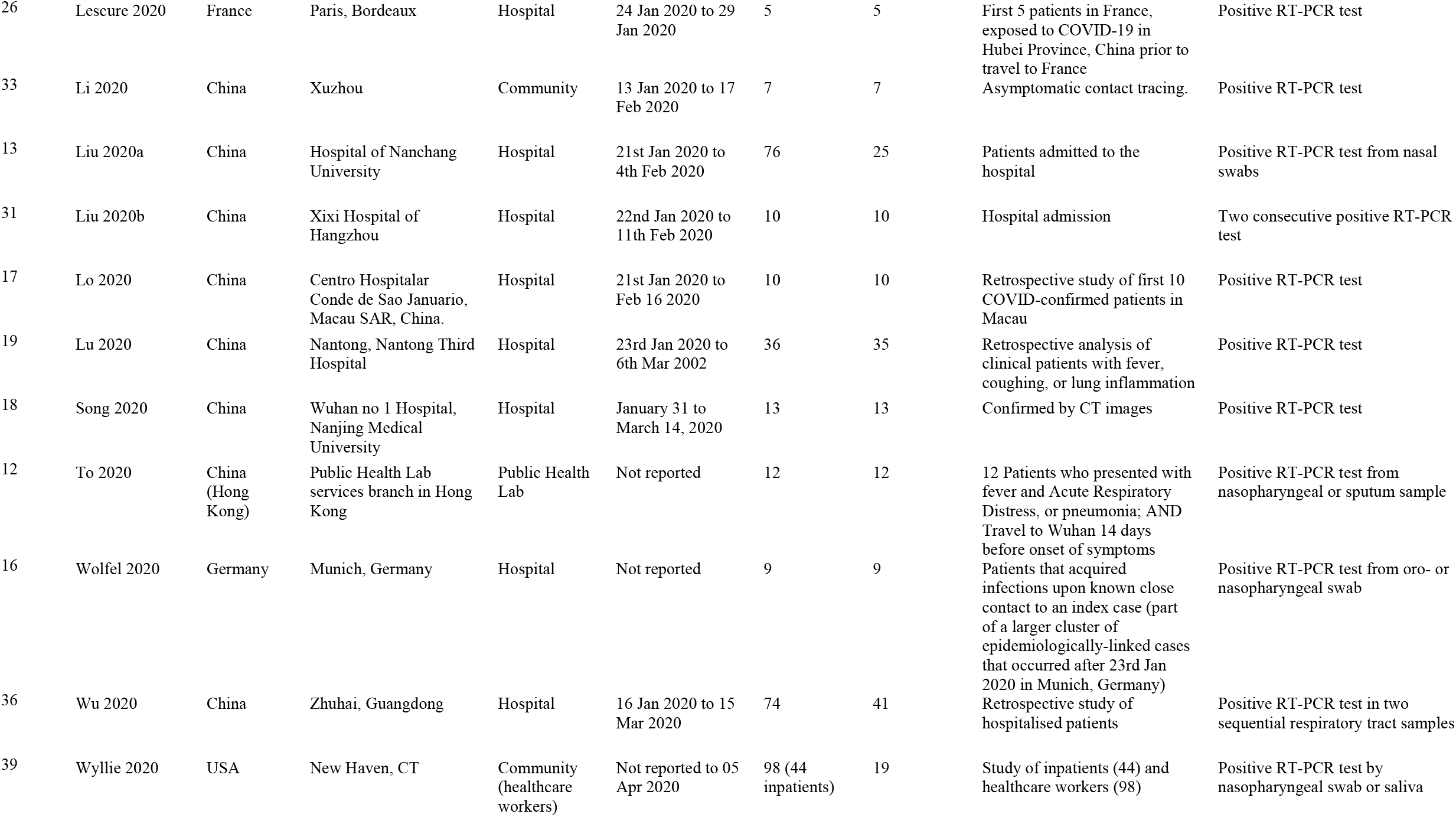
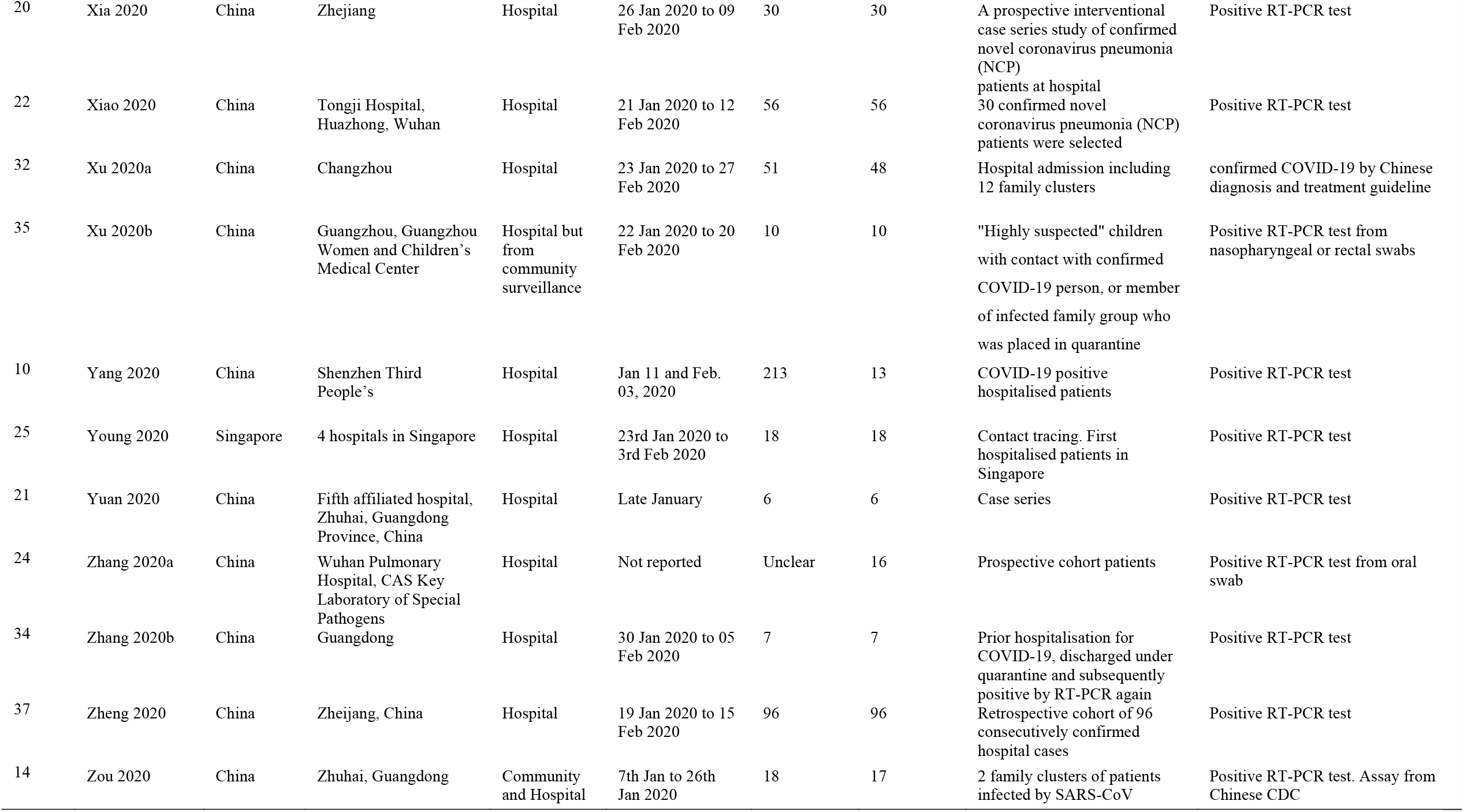
Table of study characteristics study design.

**Table 2:** Table of study characteristics IPD participant, sample site, test type.

| Ref | Author Year | How were IPD participants selected? | % male | Age median (yrs) | Age mean (yrs) | Age range (yrs) | Sample site | PCR Test Type | Name of test equipment |
| --- | --- | --- | --- | --- | --- | --- | --- | --- | --- |
| 8 | Cai 2020 | IPD of almost all patients in study | NR | NR | NR | NR | Not reported | RT-PCR | Not reported |
| 15 | Chang 2020 | IPD of all patients in study | 69 | 36 | Not reported | IQR 24-43 | Throat | qPCR | Not reported |
| 9 | Chen 2020a | Initial or follow-up positive sputum or faecal samples paired with a follow-up negative pharyngeal sample | 64 | 37 | 37 | IQR 30-49 | Faeces, Sputum, URT (Pharyngeal) | RT-qPCR | Not reported |
| 11 | Chen 2020b | IPD of all patients in study | 51 | 51 |  | IQR 36-64 | URT | RT-PCR | Not reported |
| 28 | He 2020 | IPD for all 94 patients presented, but could only extract 19 IPD from overlapping graphs | 50 | 47 |  |  | Throat | RT-PCR | not listed, N-gene specific |
| 23 | Hu 2020a | IPD of all patients in study | 48 | 46 | NR | NR | Nasopharyngeal | RT-PCR Orflab and N | Qingdao CDC |
| 29 | Hu 2020b | IPD of almost all patients in study | 33 | 33 | 38 | 5-95 | Throat | RT-PCR | BGI Genomics qRT-PCR kits, automatic nucleic acid extraction (BioPerfectus technologies) |
| 27 | Jiehao 2020 | IPD of all patients in study | 40 | 7 | 6 | 3 months - 11 years | URT | RT-PCR Orflab and N | Not reported, CDC laboratory |
| 30 | Kujawski 2020 | IPD of all patients in study | 67 | 53 |  | 21-68 | Throat, Nasopharyngeal, Sputum, Urine, Faeces | RT-PCR | unspecified, CDC approved |
| 38 | Lavezzo 2020 | All residents identified with infection | 50 | NR | NR | NR | Nasopharyngeal | RT-PCR RdRp and E genes | in-house probes, RT-PCR assay on ABI 7900HT (ThermoFisher) |
| 26 | Lescure 2020 | IPD of all patients in study | 60 | 46 | 47 | 30-80 | Nasopharyngeal, Faeces, Conjunctiva, Urine, Blood, LRT (Pleural) | RdRp-IP1 and RdRp quantitative rtRT-PCR (appendix p 4) | RNA extraction with 'Extraction NucleoSpin DX Virus Kit' or by 'NucliSENS easyMAG', Primer and Probes 'Eurogentec' for RdRp or E-gene. |
| 33 | Li 2020 | IPD of all patients in study | 57 | 42 | 43 | 21-62 | Throat | RT-PCR type not reported | Not reported, CDC laboratory |
| 13 | Liu 2020a | 25 participants with serial samples tested for PCR | NR | NA | NA | NA | Nasopharyngeal | RT-PCR | No Name Given |
| 31 | Liu 2020b | IPD of all patients in study | 40 | 42 |  | 34-50 | mixed URT(Nasal, Throat) | RT-PCR | equipment not mentioned. |
| 17 | Lo 2020 | IPD of all patients in study | 30 | 54 | NR | NR | Faeces, Nasopharyngeal, Urine | RT-PCR (ORF1ab/N gene) | BioGerm. |
| 19 | Lu 2020 | IPD of all patients in study | NR | NR | NR | NR | Sputum, URT (Pharyngeal), Faeces, Blood | compares digital PCR to RT-PCR. Compares 4 primer sets for each, but these mostly give the same result | dPCR DropX-2000, RT_PCR reported as officially approved clinical RT-PCR from local CDC |
| 18 | Song 2020 | IPD of all patients in study | 100 | NR | NR | 22-67 | URT (Pharyngeal) | RT-qPCR or virus antibody (IgM or IgG) | qRT-PCR kits (Huirui biotechnology, Shanghai, China) , IgM and IgG antibodies in serum were measured by using a commercial colloidal gold test kit (Wondfo biotechnology, Guangzhou, China) |
| 12 | To 2020 | IPD of all patients in study | 58 | 63 |  | 37 to 75 | Saliva | In house (RT-qPCR) | LightCycler 480 Real-Time PCR System (Roche), |
| 16 | Wolfel 2020 | IPD of all patients in study | NR | Not reported | Not reported | Not reported | Sputum, Faeces, URT (Pharyngeal) | RT-PCR | Superscript III one-step RT-PCR system with Platinum Taq Polymerase (Invitrogen, Darmstadt, Germany) |
| 36 | Wu 2020 | Retrospective 41 patients with positive faecal samples only | 53 | NR | NR | NR | Throat, Faeces | RT-PCR for RdRp, N and E genes | Not reported |
| 39 | Wyllie 2020 | Patients with multiple nasopharyngeal swabs or multiple saliva swabs | inpatients (52), healthcare workers(16) | NR | 61 | 23-92 | Nasopharyngeal, Saliva | RT-PCR using CDC RT-PCR primers. | NR |
| 20 | Xia 2020 | IPD of all patients in study | 70 | 51 | 55 | 13-83 | Sputum, Conjunctiva | RT-PCR | Roche Lightcycler 480 |
| 22 | Xiao 2020 | IPD of all patients in study | 61 | 55 | 55 | 25-83 | URT (Pharyngeal) | RT-PCR Orf1ab and N | Shanghai Huirui Biotechnology |
| 32 | Xu 2020a | IPD of almost all patients in study | 49 | 3 groups: imported 35[29-51], secondary 37[24-47.5], tertiary 53[35-65] |  | 24-65 | Throat | RT-PCR | Biogerm Medical Biotechnology Co., Shanghai, China |
| 35 | Xu 2020b | IPD of all patients in study | 60 | 7 | 8 | 2 months - 16 years | Nasopharyngeal, Faeces | RT-PCR Orf1ab and N | 'previously reported' |
| 10 | Yang 2020 | Not reported how 13 patients for serial sampling IPD data chosen | 51 | 52 |  | (2-86) | Mixed URT (nasal swabs, throat swabs) and mixed LRT (sputum and bronchoalveolar lavage fluid (BALF)) | RT-qPCR | QIAamp RNA Viral Kit, CFDA approved GeneoDX Co 8 detection kit |
| 25 | Young 2020 | IPD of all patients in study | 50 | 47 |  | 31-73 | Nasopharyngeal, Faeces, Urine, Blood | RT-PCR | Applied Biosystem 3500xL Genetic Analyzer |
| 21 | Yuan 2020 | IPD of all patients in study | 33 | 64 | 59 | 36-71 | Nasopharyngeal, Faeces | RT-PCR (RdRp, E and N genes) | NR |
| 24 | Zhang 2020a | Not reported | NR | NR | NR | NR | URT (Oral), Faeces | qPCR and RT-PCR spike gene | ABI 7500 machine |
| 34 | Zhang 2020b | IPD of all patients in study | 86 | 26 | 22 | 10 months - 35 years | Throat, Faeces, Blood | RT-PCR type not r+L37+L34 | Not reported |
| 37 | Zheng 2020 | IPD of all patients in study | 60 | 55 | NR | NR | Sputum, Faeces, Blood | RT-PCR for Orf1ab | BoJie |
| 14 | Zou 2020 | IPD of almost all patients in study | 50 | 59 |  | 26-76 | Throat, URT (Nasal) | RT-PCR N and Orf1b genes. Assay Chinese CDC | Not reported |

**Figure 1:**
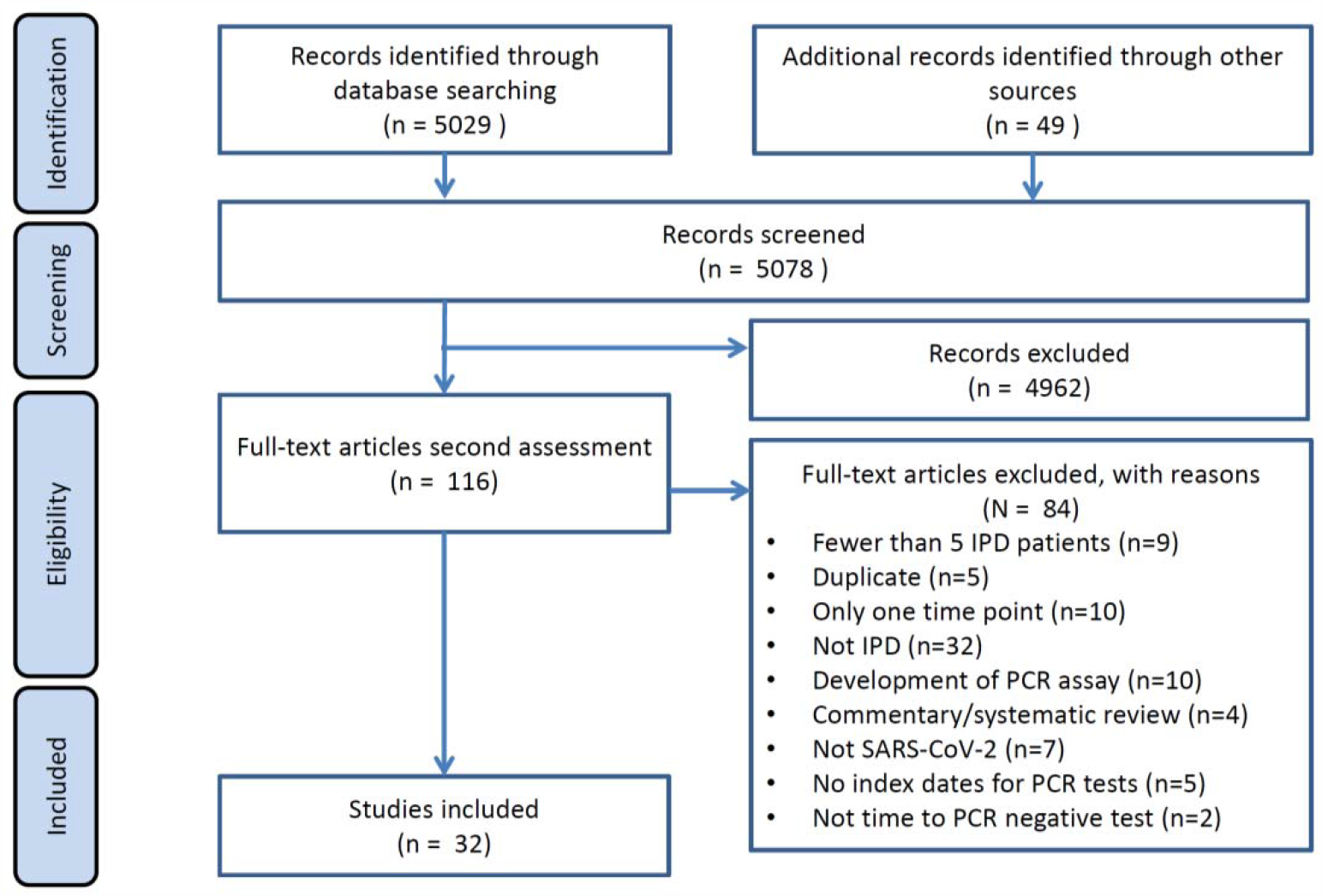
PRISMA flowchart.

### Sampling site reporting

Articles variably specified sampling sites according to anatomical location, or grouped more than one site for analysis, for example as upper RT (see S4). The most frequent sample sites were faeces (n=13), nasopharyngeal (n=10) and throat (n=9), although there was a range of other sites including blood, urine, semen, and conjunctival swabs (Table 2). Details of sampling method were generally absent. Two studies specified the person taking the samples. One study described how the nasopharynx was identified and the swab technique (length of contact time with the nasopharynx and twisting). Five studies specified samples storage and transport details.

### Sampling site positivity over time

We present RT-PCR test results for 11 different sampling sites at different times during SARS-CoV-2 infection. Figures 2 and 3 show the number of positive and negative RT-PCR results for 5-day time intervals since symptom onset and time from hospital admission, respectively.

**Figure 2:**
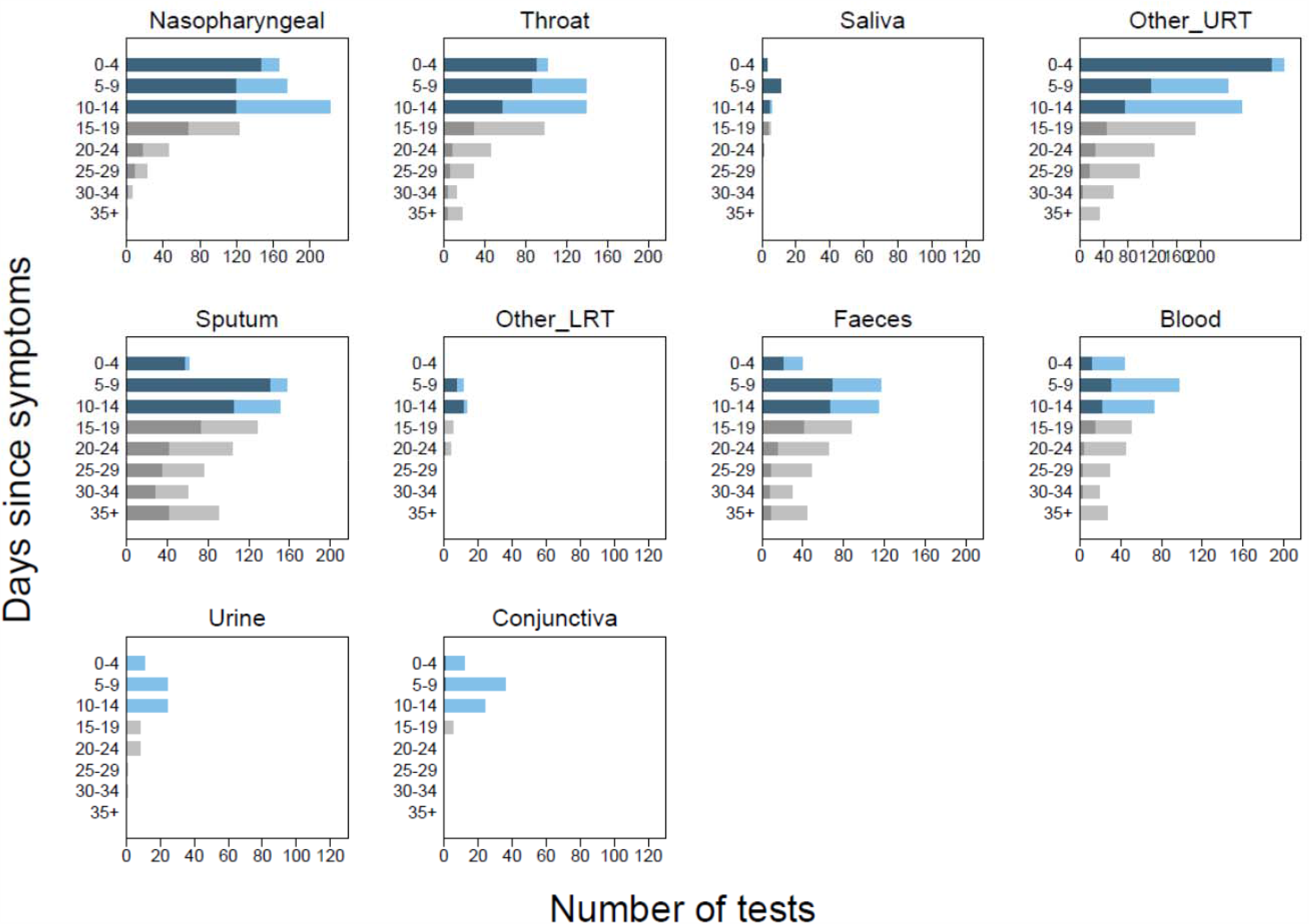
Number of positive and negative RT-PCR test results since symptom onset. Each panel shows a separate site used in participant sampling. Nasopharyngeal, saliva, sputum were used where clearly reported. Throat included throat and oropharyngeal. Other URT includes samples reported in articles as: nasal, mixed nasal and throat, oral, pharyngeal, or upper respiratory tract. For pharyngeal sampling it was not clear if this was nasopharyngeal or oropharyngeal. Other LRT includes sampling reported as lower respiratory tract or one article including pleural fluid sampling. Blood included serum, plasma, or blood. Faeces included stool or anal swab. Each panel shows 5-day time periods since the onset of symptoms: 0-4 days, 5-9 days, 10-14 days, 15-19 days, 20-25 days, 26-30 days, 31-34 days, 35 to max days. The numbers of positive RT-PCR tests are shown as dark blue bars and dark grey bars between 0 to 14 days and 15 to 40 days respectively, and the number of negative RT-PCR results are shown similarly as light blue bars and light grey bars. Different colours are used before and after 15 days to indicate caution, as after 15 days testing is enriched in more severely ill participants. The total number of tests within a particular time period can be read from the x-axis.

**Figure 3:**
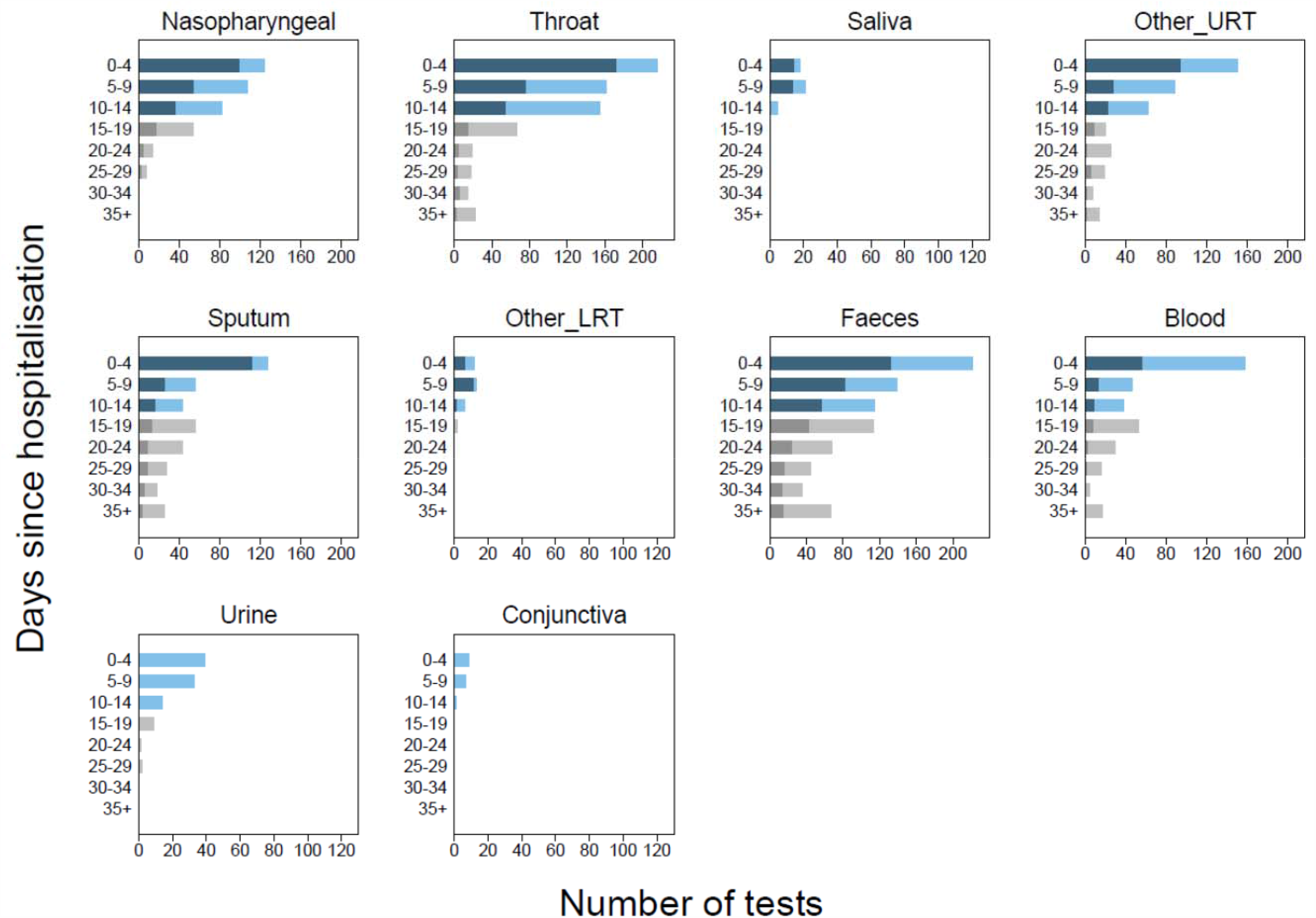
Number of positive and negative RT-PCR test results since hospital admission. Each panel shows a separate site used in participant sampling. Nasopharyngeal, saliva, sputum were used where clearly reported. Throat included throat and oropharyngeal. Other URT includes samples reported in articles as: nasal, mixed nasal and throat, oral, pharyngeal. or upper respiratory tract. For pharyngeal sampling it was not clear if this was nasopharyngeal or oropharyngeal. Other LRT includes sampling reported as lower respiratory tract or one article including pleural fluid sampling. Blood included serum, plasma, or blood. Faeces included stool or anal swab. Each panel shows 5-day time periods since the hospital admission: 0-4 days, 5-9 days, 10-14 days, 15-19 days, 20-25 days, 26-30 days, 31-34 days, 35 to max days. The numbers of positive RT-PCR tests are shown as dark blue bars and dark grey bars between 0 to 14 days and 15 to 40 days respectively, and the number of negative RT-PCR results are shown similarly as light blue bars and light grey bars. Different colours are used before and after 15 days to indicate caution, as after 15 days testing is enriched in more severely ill participants. The total number of tests within a particular time period can be read from the x-axis.

The sampling sites yielding the greatest proportion of positive tests were nasopharyngeal, throat, sputum, or faeces. Insufficient data were available to evaluate saliva and semen. Only 33% of participants who were tested with blood samples had detectable virus (44/133), and almost no samples from urine or conjunctival sampling detected virus presence.

Using nasopharyngeal sampling, 89% (147/166, 95% CI 83 to 93) RT-PCR test results were positive between 0 to 4 days post-symptom onset and 81% (100/124, 95% CI 73 to 87) 0 to 4 days post hospital admission (Figure 2 and 3). At 10 to 14 days the percentage of test results positive reduced to 54% (120/222, 95% CI 47 to 61) post-symptoms and 45% (37/82, 95% CI 34 to 57) post-admission (S6, S7).

Using throat sampling at 0 to 4 days post-symptoms, 90% (91/101, 95% CI 83 to 95) of test results from participants with SARS-CoV-2 were detected by RT-PCR sampling, falling to 42% (58/139, 95% CI 33 to 50) at 10 to 14 days post-symptom onset (Figure 2, S6, S7). Similar results were observed for time since hospital admission, where at 0 to 4 days 80% (173/215, 95% CI 75 to 86) of result were positive, falling to 35% (55/155, 95% CI 28 to 44) between 10 to 14 days (Figure 3, S6, S7). Using faecal sampling, 55% test results are positive (22/40, 95% CI 38 to 71) at 0 to 4 days post-symptom onset.

### Upper and lower respiratory tract sampling

We further grouped sites into upper (URT) and lower (LRT) respiratory tract. The rate of sample positivity reduced faster from URT sites compared to LRT sites (Figure 4A). Given that analysis across all participants is likely to be influenced by preferential URT sampling of participants with less severe disease, we also analysed participants who underwent both URT and LRT sampling. Again, URT sites on average cleared faster (median 12 days, 95% CI 8 to 15 days) than LRT sites (median 28 days, 95% CI 20 to not estimable; Figure 4B); the majority of participants clear virus from URT site before LRT (Figure 4C). Data based on time since hospital admission are consistent with data for time since symptom onset.

**Figure 4:**
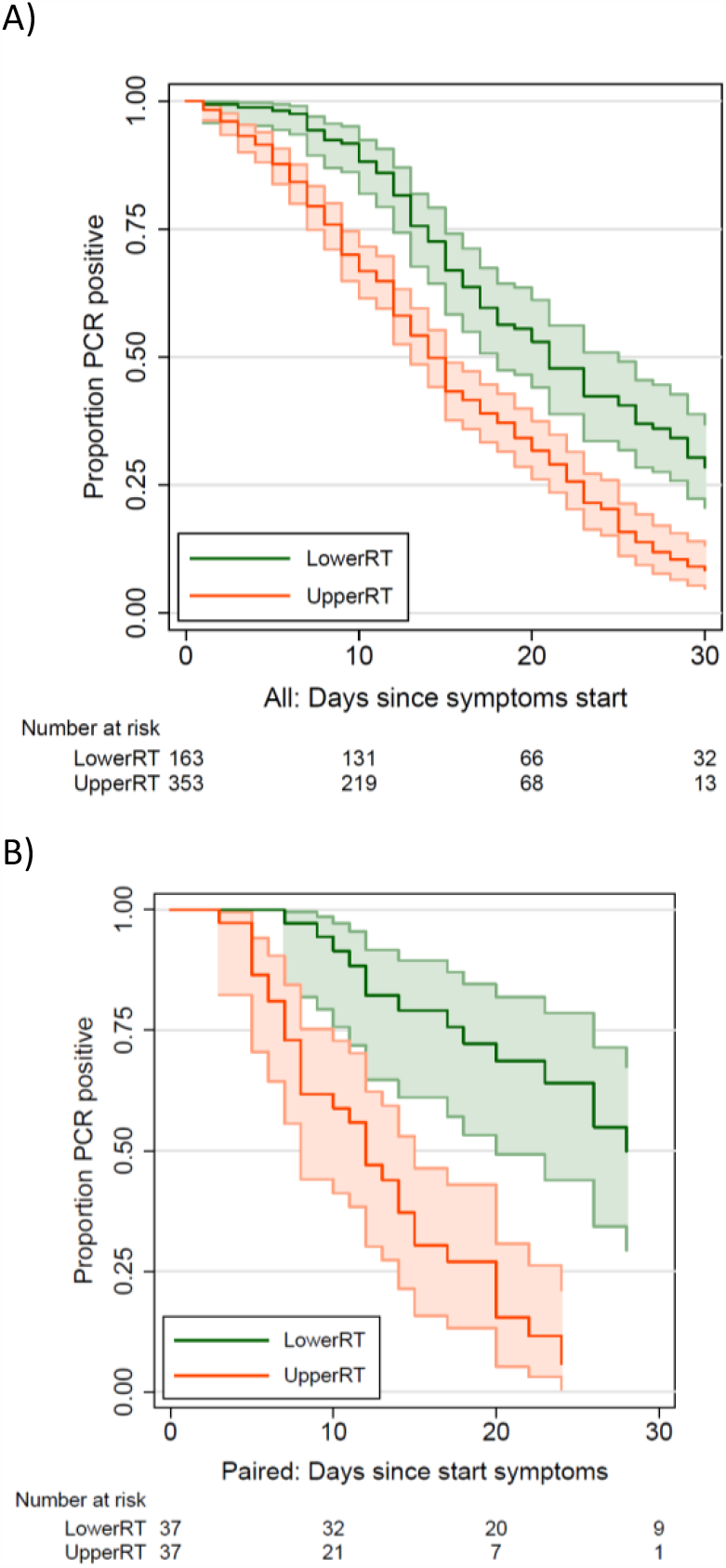
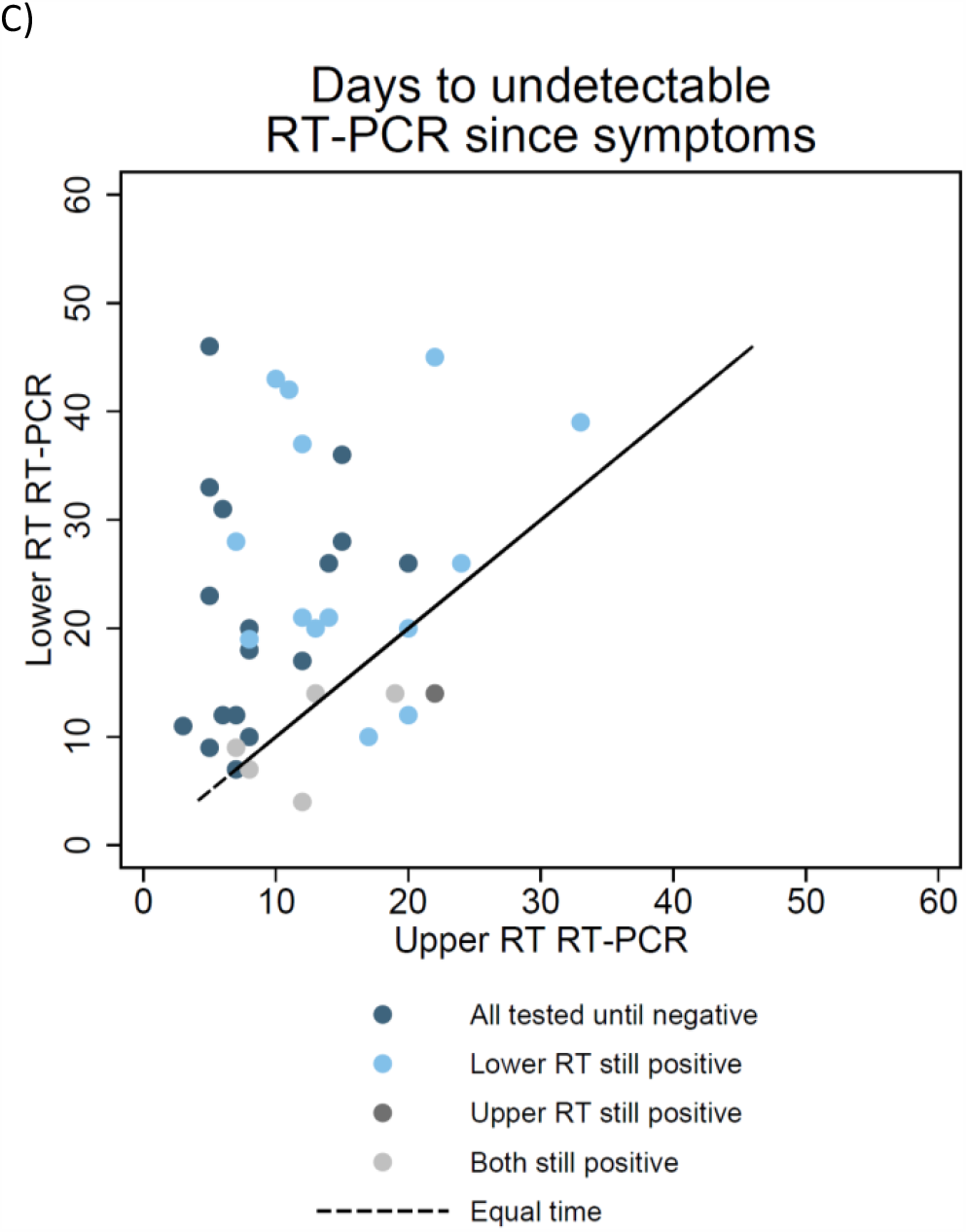
Comparison of duration of detectable virus from upper and lower respiratory tract sampling. (A) Time to undetectable virus in upper and lower respiratory tract samples. Kaplan-Meier with 95% confidence intervals and number at risk. All samples in review. (B) Time to undetectable virus in upper and lower respiratory tract samples in participants who were tested with both upper and lower respiratory tract sampling. Kaplan-Meier with 95% confidence intervals and number at risk. Restricted to participants with both sampling methods. (C) Time to undetectable virus in upper and lower respiratory tract samples in participants who were tested with both upper and lower respiratory tract sampling. Scatterplot where each dot represents a single participant, with the time to undetectable virus with both upper and lower respiratory tract sampling shown for each participant.

### Faecal vs respiratory tract sampling

Across participants sampled by both RT and faecal sampling since hospital admission, 29% of participants were detected using RT sampling but not by faecal sampling, (52/177 participants, 95% CI 23 to 37%, 10 studies). The time to RT-PCR tests becoming undetectable varied greatly by participant, although time to undetectable virus was similar for both sampling sites (Figure 5), in participants with RT-PCR test results from both RT and faecal samples. Thirty nine out of 89 participants (44%, 95% CI 33 to 55%) had a shorter duration of detection in faecal samples than in RT samples.

**Figure 5:**
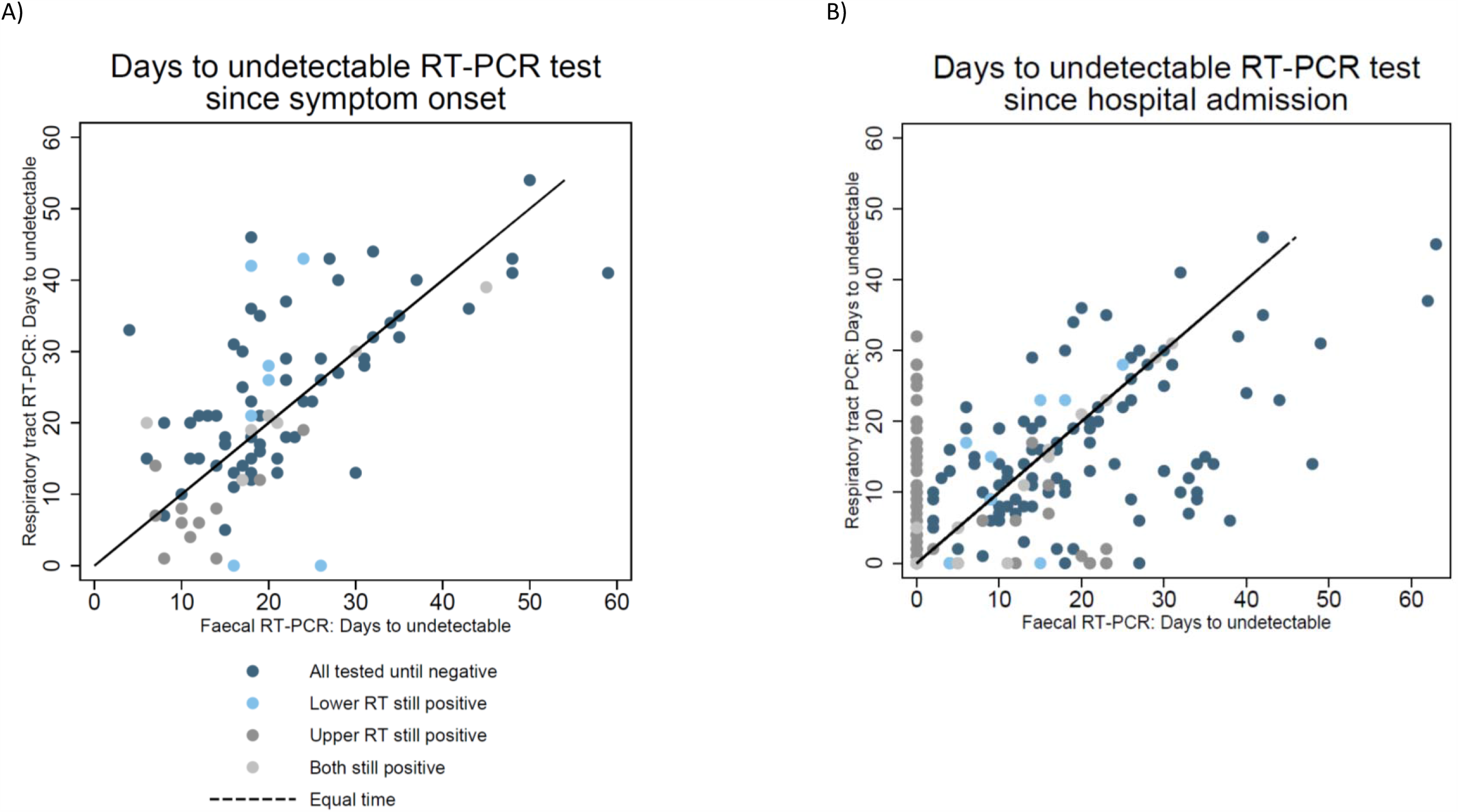
Comparison of days to undetectable virus from respiratory tract and faecal sampling. Time to undetectable virus in faecal compared to any respiratory tract sample in participants who were tested with both sampling. Scatterplot where each dot represents a single participant, with the time to undetectable virus with both faecal and respiratory tract sampling shown for each participant. Thirty percent of participants tested at both sampling sites do not have detectable virus in faecal samples.

Median time to clearance from RT was shorter in participants based on time since hospitalisation (125 participants, p=0.014), whilst similar in participants since onset of symptoms (87 participants, p=0.15) (S8).

### Intermittent false negative results

Many articles reported intermittent false negative RT-PCR test results for participants within the monitoring time span. Where participant viral loads were reported, several different profiles were distinguished; two examples are shown in Figure 6^13,14^. Intermittent false negative results were reported either where the level of virus is close to the limit of detection, or in participants with high viral load but for unclear reasons.

**Figure 6:**
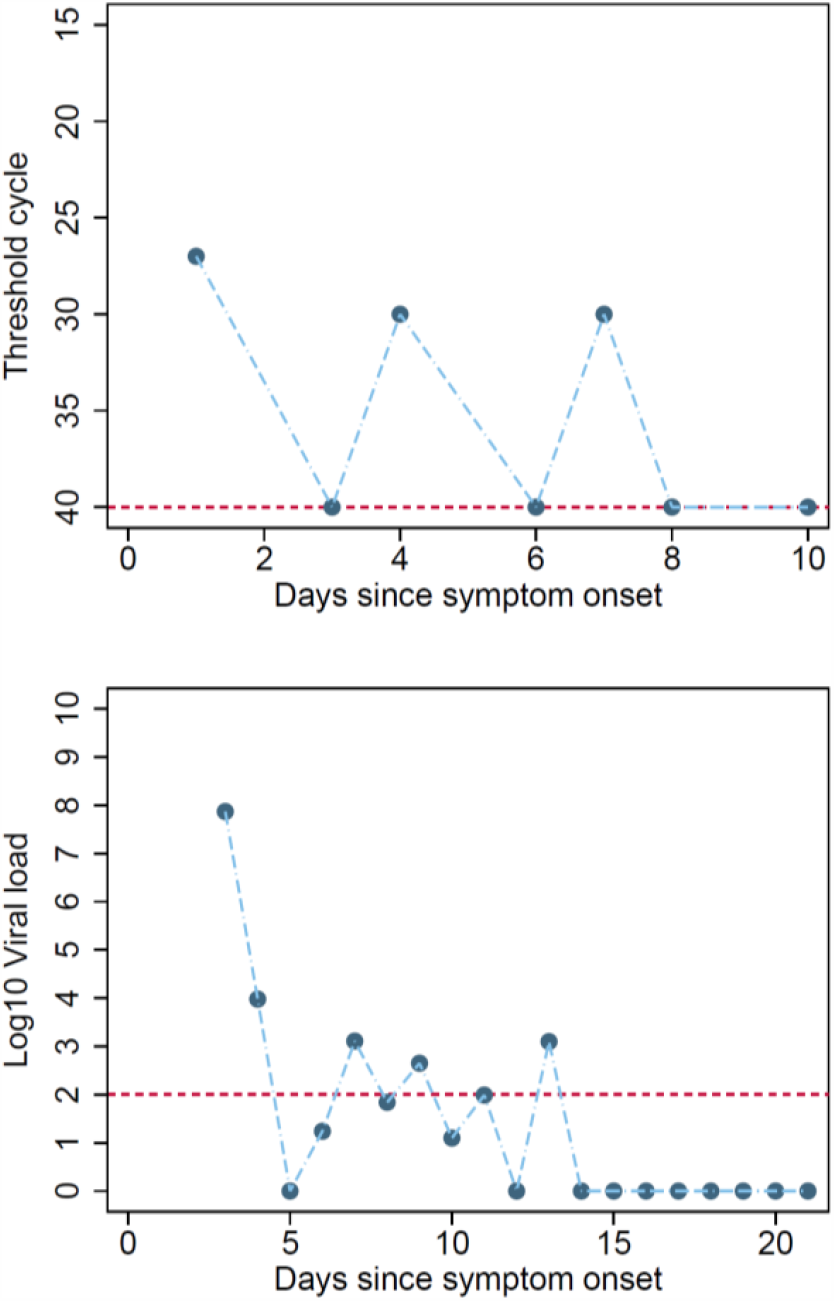
Example participants with intermittent false negative results. Figure 6A show a participant where virus levels have reduced over time to a level around the limit of viral detection, and at these low levels of virus intermittent negative results will occur due to differences in the location or amount of sample. Figure 6B shows an example of a participant with high viral load, but where alternate RT-PCR test results report high viral load or undetectable virus.

### Risk of bias

The proportion of studies with high, low, or unclear ROB for each domain is shown in Figure 7 and ROB for individual studies is shown in S5. All studies were judged at high ROB. All but one were judged at high ROB for the participant selection domain^16^, mainly as they only included participants with confirmed SARS-CoV-2 infection based on at least one positive PCR test. Studies also frequently selected a subset of the participant cohort for longitudinal RT-PCR testing, and only results for these participants were included in the study. Ten studies were judged at unclear ROB for the index test domain as the schedule of testing was based on clinician choice rather than being pre-specified by the study or clinical guidelines, or because the samples used for PCR testing were not pre-specified. Eleven studies were judged at high ROB for the flow and timing domain mainly because continued testing was influenced by easy access to participants, such as by continued hospitalisation.

**Figure 7:**
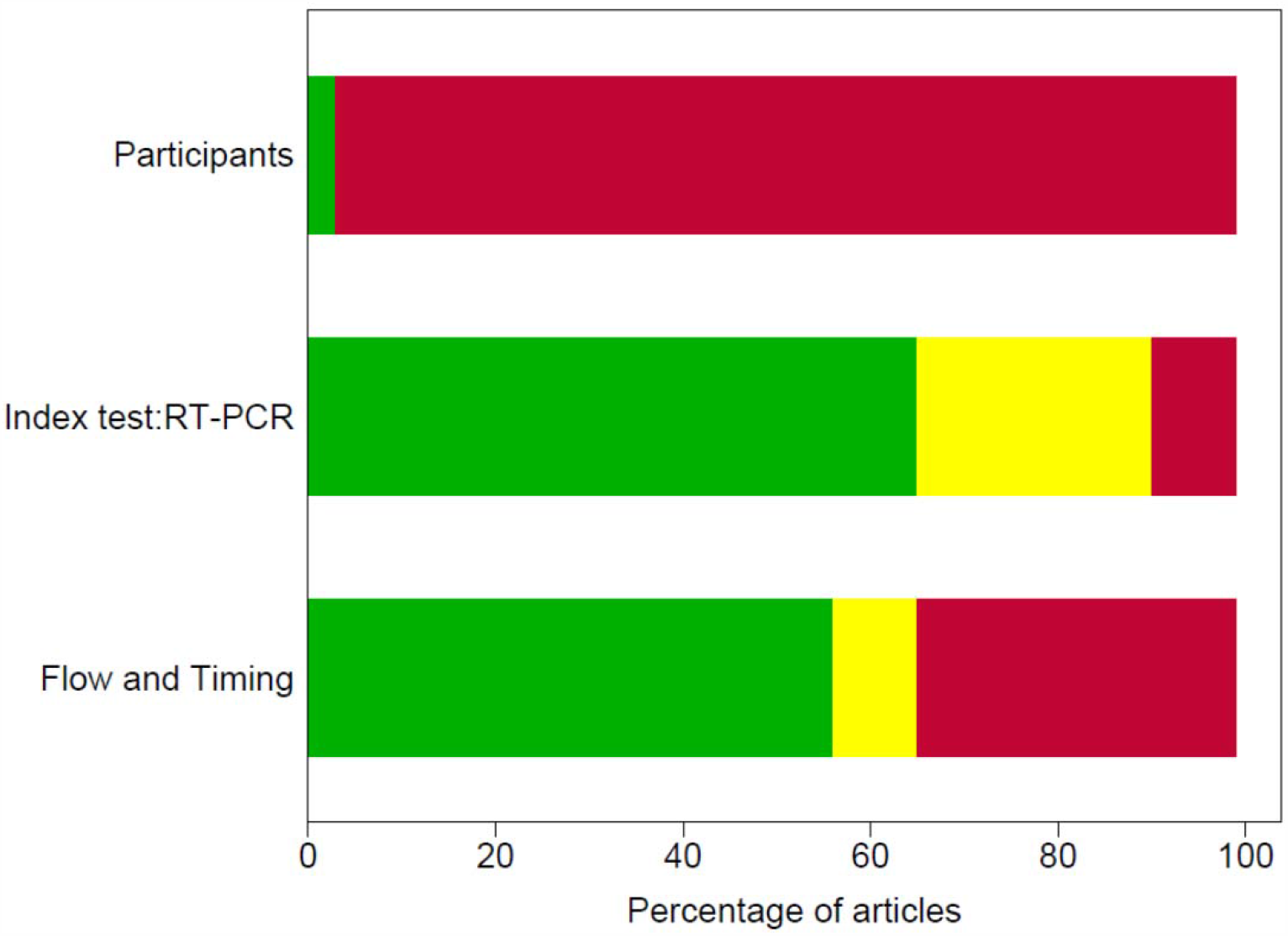
Risk of bias by adapted QUADAS-2 domain. An adapted version of QUADAS-2 for longitudinal studies was used (Supp_table 3). For each domain, the percentage of studies by concern for potential risk of bias is shown: low (green), unclear (yellow) and high (red).

## DISCUSSION

### Key findings

Negative RT-PCR test results were common in people with SARS-CoV-2 infection confirming that RT-PCR testing misses identification of people with disease. Our IPD systematic review has established that sampling site and time of testing are key determinants of whether SARS-CoV-2 infected individuals are identified by RT-PCR.

We found that nasopharyngeal sampling was positive in approximately 89% (95% CI 83 to 93) of tests within 4 days of either symptom onset. Sampling ten days after symptom onset greatly reduced the chance of a positive test result.

There were limited data on new methods of sample collection like saliva in these longitudinal studies. Sputum samples have similar or higher levels of detection to nasopharyngeal sampling, although this may be influenced by preferential sputum sampling in severely ill participants. Although based on few participants tested at both sampling sites, URT sites have faster viral clearance than LRT in most of these participants; 50% of participants were undetectable at URT sites 12 days after symptom onset compared to 28 days for LRT.

We found that faecal sampling is not suitable for initial detection of disease, as up to 30% of participants detected using respiratory sampling are not detected using faecal sampling. Viral detection in faecal samples maybe useful to establish virus clearance, although as noted, whether RT or faecal samples have longer duration of viral detection varies between participants.

All included studies were judged at high ROB so results of this review should be interpreted with caution. Box 1 provides an overview of the major methodological limitations and their potential impact on study results. A major source of bias is that all but one study ^18^ restricted inclusion to participants with confirmed SARS-CoV-2 infection based on at least one positive RT-PCR test, meaning that the percentage of positive RT-PCR testing is likely to be overestimated.

Lack of technical details, for example of how samples are taken and RT-PCR tests performed, limit the applicability of findings to current testing. Compared to real life, studies were likely to use more invasive sampling methods, use experienced staff to obtain samples, and sample participants in hospital settings where sample handling could be standardised. Consequently, estimates of test performance are likely to be overestimated compared to real-world clinical use and in community population testing including self-test kits.

These limitations have important implications for how testing strategies should be implemented and in particular how a negative RT-PCR test result should be interpreted.

### Putting the findings into context of literature

The accuracy of RT-PCR testing is limited by sampling sites used, methods and the need to test as soon as possible from symptom onset in order to detect the virus. Previous studies have established that in COVID-19 infection viral loads typically peak just before symptoms and at symptom onset^3^. To our knowledge, there has been no prior systematic review of RT-PCR using IPD to quantify the percentage of persons tested who are positive and how this varies by time and sampling site.

Understanding the distribution of anatomical sites with detectable virus is clinically relevant, especially given independent viral replication sites in nose and throat using distinct and separate genetic colonies^16^. Understanding of different patterns of detection and duration of virus detection at different body sites is essential when designing strategies of testing to contain virus spread. Notably, it is unclear if detection of virus in faeces is important in disease transmission, although faecal infection was shown in SARS and MERS^40^.

### Strengths of study

This review uses robust systematic review methods to synthesise published literature and identifies overall patterns not possible from individual articles. Using IPD, we examined data across studies and avoided study level ecological biases present when using overall study estimates. IPD regarding sample site at different timepoints during infection is vital because it provides an overview of test performance impossible from individual studies alone. Synthesised IPD can also substantiate or reject patterns appearing within individual studies. Within participant paired comparisons of sampling sites also becomes possible with sufficient data.

### Limitations of study

The main limitation is the risk of bias in the included studies. Although constraints were understandable given the circumstances in which the studies were done, the consequences for validity need to be highlighted. The percentage of positive RT-PCR testing is likely to be overestimated, because inclusion was restricted to participants with confirmed SARS-CoV-2 infection based on at least one positive RT-PCR test in all but one study^18^. This means that people who had a COVID-19 infection but never tested positive on at least one RT-PCR test would not have been included. This could arise if SARS-CoV-2 is not present at easily sampled sites or at the time participants were tested. This makes it impossible to determine the true false negative rate of the test – the proportion of people who actually have SARS-CoV-2 but would receive a negative RT-PCR test result. It is possible that only half of persons infected by SARS-CoV-2 may test positive, as a community surveillance study in Italy found only 53% (80/152) persons tested RT-PCR positive in households quarantined for 18 days with persons who tested PCR positive^38^. The same study also identified households where no one tested RT-PCR positive, but where there were clusters of persons with symptoms typical of COVID.

Poor reporting of sampling methods and sites impaired our ability to distinguish between and report on variability between them. For some sampling methods such as saliva and throat swabs, more studies are needed. There were also sparse data on sampling methods that are becoming more widespread, such as participant self-sampling^41^ and short nasal swab sampling (anterior nares/mid turbinate)^42^. Our index times may be subject to bias as symptom onset is somewhat subjective and hospital admission practices vary by country, pandemic stage, and hospital role (i.e. healthcare vs. isolation). The results presented do not correspond to following the same participants across time, but the testing at clinically relevant time snapshots reported from individual studies, so that participants tested at later time points are likely to have more severe disease; this does not limit the interpretation of results in understanding testing of participants in most clinical contexts. Comparisons of sampling sites should be restricted to participants tested at the relevant sites. We have used analysis methods that do not include clustering within studies, to keep analyses simple to understand and present, and to avoid complications of fitting models where the number of participants in each cluster varies. Ultimately, many potentially eligible studies did not report IPD which led to their exclusion, or only reported IPD for a subset of participants in the study. We would welcome contact and data sharing with clinicians and authors to rectify this.

### Implications for policy/practice/future research

To avoid the consequences of missed infection, samples for RT-PCR testing need to be taken as soon as symptoms start for detection of SARS-CoV-2 infection in preventing ongoing transmission.

Even within four days of symptom onset some participants infected with SARS-CoV-2 will receive negative test results. Testing at later times will result in a higher percentage of false negative tests in people with SARS-CoV-2, particularly at upper RT sampling sites. After ten days post-symptoms it may be important to use lower RT or faecal sampling. Valid estimates are essential for clinicians interpreting RT-PCR results. However, ROB considerations suggest that the positive percentage rates we have estimated may be optimistic, possibly considerably so.

Participants can have detectable virus in different body compartments, so virus may not be detected if samples are only taken from a single site. Some hospitals in the United Kingdom now routinely take RT-PCR samples from multiple sites, such as the nose and throat. More studies are urgently needed on evolving sampling strategies such as self-collected samples which include saliva and short nasal swabs. Future studies should avoid the risks of bias we have identified by precisely reporting the anatomical sampling sites with a detailed methodology on sample collection.

Further sharing of IPD will be important and we would welcome contact from groups with IPD data we can include in ongoing research.

## Data Availability

Data is available from original publications

## CONTRIBUTOR STATEMENT

SM conceived the study and design.

JA, SG, BS, PT, CH, ST, NS, PW, and RW contributed to study design.

NR, SM, BS, ZZ, CH, JP, Virus Bashers team, SG, and LFdR selected the articles.

SM, BS, ZZ, CH, JP, KG, JS, JA, SG, and AW extracted the data.

SM analysed the data.

SM, SH, ST, NS, CH, PW and RW wrote the first and second drafts of the manuscript.

SM, SH, ST, NS, CH, PW, RW, JA, SG, KG, JS, BS, ZZ, JP, PT, NR, LFdR, GB, BN, and AW interpreted the data and contributed to the writing of the final version of the manuscript.

All authors agreed with the results and conclusions of this article.

## DECLARATION OF INTERESTS

SM, SH, AJA, SG, KG, SAT, BS, JP, PT, PW, JS, LFR, NR, NS, JS, AW, JA, GB, KG, BN declare no conflicts of interest for the submitted work.

CH has advised Attomarker, a spin-out company of the University of Exeter about the conduct of evaluations of its tests for COVID antibodies, but received no payment for this advice and provided it as part of academic duties at the University of Exeter

## FUNDING

No specific funding has been received for this research.

SM is funded on grant funding from the National Institute for Health Research (NIHR),

University College London (UCL) and UCL Hospital Biomedical Research Centre.

SAT is an NIHR senior investigator.

NS receives funding from the UCL/UCLH Biomedical Research Centre.

AJA, SG, KG, JS and AW are funded by the NIHR Newcastle In Vitro Diagnostics Co-operative.

BS is part-funded by NIHR Leeds In Vitro Diagnostic Co-operative.

PT is funding by NIHR MedTech and In-vitro Diagnostics Co-operative at Oxford University LFR is funded by a UK MRC Methodology Programme Grant (No. MR/T025328/1).

BDN is an NIHR Academic Clinical Lecturer

## ACKNOWLEDGEMENTS

**“Virus Bashers” team**

Steve Harris, Christine Bennett, Tim Carne, Chris Challis, Christian Dixon, Jan Dixon, John Golightly, Sue Hamer-Moss, Claire Hicks, Brenda Johnston, Lionel Murphy, Lois Norton, Jonathan Sanvoisin, Glen Titmus, Stuart Wilson & Friends

## ABBREVIATIONS

BAL: bronchoalveolar lavage
BALF: bronchoalveolar lavage fluid
CI: confidence interval
Ct: cycle threshold
HIV: human immunodeficiency virus
IPD: individual participant data
KM: Kaplan-Meier
LRT: lower respiratory tract
MERS: Middle East respiratory syndrome
MeSH: Medical Subject Headings
NIHR: National Institute for Health Research
QUADAS-2: A Revised Tool for the Quality Assessment of Diagnostic Accuracy Studies
RNA: ribonucleic acid
ROB: Risk of Bias
RT-PCR: reverse transcriptase polymerase chain reaction
RT: respiratory tract
SARS: Severe acute respiratory syndrome
TOC: Table of Characteristics
UK: United Kingdom
URT: upper respiratory tract
WHO: World Health Organization

## FIGURES AND TABLES

**BOX 1:** Biases and issues in interpretation

BOX 1: Biases and issues in interpretation
| Domain | Details of bias and applicability issues | Impact on interpretation of study data |
| --- | --- | --- |
| **Participants (source of bias)** | In these studies, the reference test usually incorporates RT-PCR (index test)<br>RT-PCR testing is usually a key component of identifying people with SARS-CoV-2 infection<br>Participants will not be detected or included in these studies when SARS-CoV-2 is not present at easily sampled sites and at the time that participants were available for testing | Unclear how many and what severity of participants with SARS-CoV-2 are not included in studies<br>People who do not have a positive RT-PCR test at some point are excluded. This could lead to over-estimation of positivity<br>Rates of positivity will be inflated as only people with virus accessible for sampling for RT-PCR tests will be included in studies |
| **Participants (source of bias)** | Most participants are identified or present based on respiratory tract symptoms such as cough or respiratory distress.<br>Participants will not be detected or included in these studies when less common symptoms or asymptomatic<br>Participants included will be biased to over-represent people with detectable virus in respiratory tract sampling sites and at times frequently used for testing (post symptom onset or at admission to hospital) | Unclear how many and what severity of participants with SARS-CoV-2 are not included in studies<br>Studies will inflate positivity for sampling sites that overlap with sampling sites used in RT-PCR reference testing<br>For example, we identified 30% of participants with RT positivity but with negative results from faecal sampling. However, if participants had only faecal virus, would they have been included in the studies? |
| **Index test: RT-PCR (applicability)** | Studies included are likely to use more invasive sampling methods than acceptable in widespread population testing. For example, nasopharyngeal testing is likely in many current studies to be based on long swabs and self test kits<br>Studies will use experienced staff to obtain samples, handle, process and conduct tests<br>Studies are mostly sampling participants in hospital settings or in specialised research community testing research where sample handling, transport and storage have been standardised | Percentage of people with detectable virus may be overestimated when testing is applied in real world clinical use and in population testing |
|  | Variation in RT-PCR kits are minimised as studies are based in few hospitals or limited to a research setting |  |
| **Index test: RT-PCR (applicability)** | Sample RNA extraction methods are designed predominantly for respiratory samples<br>RT-PCR sample preparation kits used are mostly designed for extraction of virus from respiratory samples, not from faecal samples. It is unclear how efficiently these kits extract virus RNA from stool samples | Percentage of people with virus present in faecal samples and duration of shedding in faecal samples may be underestimated |
| **Index test: RT-PCR (applicability)** | RT-PCR tests detect both infectious and inactive (inactive due to immune system or dead) virus.<br>Few studies link RT-PCR testing to cell assays to test for infectious virus | Percentage of people with clinically important detectable virus may be overestimated |
| **Index test: RT-PCR (applicability)** | Rate of virus aggregation or clearance by immune system may affect the sampling efficiency at some sampling sites<br>Both the innate and adaptive immune system will aggregate and clear virus particles from bodily fluids. It is not clear what the time scale of clearance or how this influences detection of virus at different sites and at which time points | Percentage of people with detectable virus may be underestimated |
| **Index test: RT-PCR (applicability)** | Reporting of sampling sites and methods is poor<br>Poor reporting may have led to less ideal grouping of sampling in analysis<br>Some studies are likely to use a variety of nasopharyngeal sampling methods depending on the individual participants, but the type of sampling is typically reported at a study level for a particular sampling site | Percentage of people with detectable virus may be over or underestimated |
| **Flow and timing** | Uncertainty and inconsistencies in time of sampling<br>Time of symptom onset can be subjective unless based on fever, but some participants do not have fever.<br>Time of symptom onset may be different if asked of participants in ICU setting<br>Time of hospitalisation and discharge may be affected by function hospitalisation serves in containment of disease spread. In some studies, the hospitals were also quarantine centres, so participants were hospitalised immediately at onset of mild symptoms rather than restricted to patients needing oxygen. | Percentage of people with detectable virus may be over or underestimated at particular times |
| **Flow and timing** | Clinical cohort within studies changes across time points | Percentage of people with detectable virus may be overestimated at particular later time points as these correspond to participants who were severely ill |
|  | Participants who have recovered from COVID-19 in most studies are typically not tested after 2 negative tests 24 hours apart.<br>Many studies only test inpatients at the hospital, so the participants sampled between 0 to 14 days typically have less severe disease than those tested longer |  |
| **Flow and timing (Selective outcome reporting)** | Some studies only publish IPD data for a selection of people | Available IPD data may not represent a typical spectrum of participants in the different settings (community setting, hospital, ICU, nursing home, prison) |
| **Publication bias** | Published data is likely to be biased towards publication of research active groups which may not represent typical real world | Percentage of people with detectable virus may be overestimated |
| BAL = bronchoalveolar lavage; COVID-19 = coronavirus disease 2019; ICU = intensive care unit; IPD = individual participant data; RNA = ribonucleic acid; RT-PCR reverse transcription polymerase chain reaction; SARS-CoV-2 = severe acute respiratory syndrome coronavirus 2 |  |  |

**Box 2:** Table of examples from included studies

**Box 2: Table of examples from included studies**
| Study characteristic | Detail | Reference |
| --- | --- | --- |
| Study design: representative recruitment | Representative participants<br>Community infection<br>Contact tracing including asymptomatic<br>Hospitalised patients | Population surveillance of Italian town, with PCR testing across<sup>38</sup><br>Contact tracing<sup>8,29,33</sup><br>Retrospective cohort of 96 hospitalised patients<sup>37</sup> |
| Study design: planned testing and follow up | Informative sampling<br>Multiple sampling sites per participant<br>Planned schedule of sampling<br>Sampling continues after negative test results<br>Sampling continues after hospital discharge | Three samples per patient, multiple testing including prolonged testing even after multiple negative results<sup>9</sup><br>Population surveillance of Italian town over 18 days<sup>38</sup><br>Long follow up post hospital<sup>15,34,35</sup><br>Planned schedule of testing (<sup>29</sup> asymptomatic contact tracing followup<sup>35-37</sup>) |
| Study design: sampling | Reporting of sampling methods (sample site, staff conducting test, sample volumes & methods) | Most studies had very sparse reporting of sample collection methods.<br>Example of fuller reporting<sup>39</sup> |
| Individual participant data reporting for sharing | Examples of plots and tables that facilitated sharing of individual participant data | Retrospective cohort of 96 patients all tested with sputum, faeces, blood. Plot shows time span of positive test results, hospitalisation timing, disease severity for individual patients<sup>37</sup><br>Data showing time course of illness with PCR test results<sup>8,30</sup><br>Data showing RT-PCR test results by patient and time point<sup>9</sup><br>Viral load over time<sup>14</sup> |

